# Monitoring SARS-CoV-2 genome evolution in a localized population

**DOI:** 10.1101/2022.01.19.22269572

**Authors:** Asmita Gupta, Reelina Basu, Murali Dharan Bashyam

**Affiliations:** Laboratory of Molecular Oncology, Centre of DNA Fingerprinting and Diagnostics, Hyderabad, India

## Abstract

Despite seminal advances towards understanding its infection mechanism, SARS-CoV-2 continues to cause significant morbidity and mortality worldwide. Though mass immunization programs have been implemented in several countries, the viral transmission cycle has shown a continuous progression in the form of multiple waves. A constant change in the frequencies of dominant viral lineages, arising from the accumulation of nucleotide variations (NVs) through favourable selection, is understandably expected to be a major determinant of disease severity and possible vaccine escape. Indeed, worldwide efforts have been initiated to identify specific virus lineage(s) and/or NVs that may cause a severe clinical presentation or facilitate vaccination breakthrough. Since host genetics is expected to play a major role in shaping virus evolution, it is imperative to study role of genome-wide SARS-CoV-2 NVs across various populations. In the current study, we analysed the whole genome sequence of 3543 SARS-CoV-2 infected samples obtained from the state of Telangana, India (including 210 from our previous study), collected over an extended period from April, 2020 to October, 2021. We present a unique perspective on the evolution of prevalent virus lineages and NVs during this time period. We also highlight presence of specific NVs likely to be associated favourably with samples classified as vaccination breakthroughs. Finally, we report genome-wide intra-host variations (iSNVs) at novel genomic positions. The results presented here provide critical insights into virus evolution over an extended time period within a geographically restricted area and pave the way to rigorously investigate the role of specific NVs in vaccination breakthroughs.

## Introduction

In December 2019, a local outbreak of multiple cases of acute pneumonia (later classified as coronavirus disease (COVID-19, https://www.who.int/news-room/q-a-detail/coronavirus-disease-covid-19)) was reported in Wuhan, Hubei province, China, caused by a novel coronavirus called as Severe Acute Respiratory Syndrome coronavirus 2 (SARS-CoV-2). The outbreak was soon followed by a rapid worldwide transmission which led to World Health Organization (WHO) declaring COVID-19 as a global pandemic in January, 2020. The rapid transmission rate of SARS-CoV-2 has resulted in 281,808,270 infections and >5 million deaths worldwide (as per WHO statistics, collected till December 29, 2021). More importantly, the last one year witnessed the emergence of multiple virus variants[1,2], of which five were classified as Variants of Concern (VoCs) by the WHO (https://www.who.int/en/activities/tracking-SARS-CoV-2-variants/) based on their higher transmissibility[3–6] and/or enhanced ability to escape neutralization by antibodies[7–12]. Of the five, the B.1.617.2 lineage (designated (and hereinafter mentioned) as ‘Delta’ by the WHO) has exhibited maximum transmission during the past one year making it the predominant viral form present worldwide[13]. As SARS-CoV-2 continues to evolve into distinct lineages, with potentially increased pathogenicity and/or transmission abilities, it becomes imperative to study the emerging genomic variants.

The current study was initiated to analyse the sequences generated from SARS-CoV-2 infected samples, collected over a course of 19 months from the state of Telangana, India. We present the major trends of dominant lineages propagating in Telangana and corresponding trends in other pan-Indian regions and worldwide, across this time period. We also report a comprehensive map of all nucleotide variations (NVs) in the viral genome during this period and their possible association with vaccine escape. Finally, we have analysed specific virus genomic positions involved in generating intra-host diversity.

## Materials and Methods

### Sample collection strategy, dataset structure and features

A total of 3543 samples (1407 females and 2091 males (information unavailable for 45 samples)), representing the period April 1, 2020 to October 31, 2021, and belonging to Telangana, India, were analysed in this study (Table S1A). The sample collection strategy for the period April, 2020 till February, 2021 was unchanged from our previous study[14]. For the subsequent period, Nasopharyngeal/Oropharyngeal swabs were collected from several RT-PCR based testing centres as well as multi-speciality hospitals across Telangana, as per guidelines established by the Indian SARS-CoV-2 Genome Consortium (INSACOG)[15]. In addition, samples received in the Covid-19 testing laboratory in CDFD, Hyderabad, were also included in the study. The work was initiated following approvals from the Institutional Bioethics committee and Biosafety committee. The sample collection peaked during the months of June and July, 2020, followed by a hiatus, and subsequently increased from March, 2021 onwards, roughly coinciding with the first and second waves of the pandemic, respectively. Of the total cases, 360 represented the age group <18 years, 2674 represented age group 18-60 years, 355 represented the age group > 60 years, while age was not documented for 154 cases. The dataset also comprised of two independent sets of cases belonging to local isolated transmission clusters (so called ‘super-spreader’ events) (Table S1A).

Cases were classified as vaccination breakthroughs if they reported infection ≥14 days of receiving second dose of either ChAdOx1 or BBV152 vaccine or ≥21 days of receiving first dose[16],[17]. The dataset included a total of 313 vaccination breakthrough cases, of which 244 belonged to Telangana, India, 18 to Uttar Pradesh, India (obtained from the Banaras Hindu University), and 51 to Chennai, Tamil Nadu, India (obtained from the Department of Public Health and Preventive Medicine, State Public Health Laboratory) (Tables S1B, S1C). Of these, 154 were completely vaccinated, 149 had received only one vaccination dose, while the status of 10 was unavailable. The majority of cases (228/313; 72.8%) received the ChAdOx1 nCov-19 vaccine^18^ (commercial name – Covishield) developed at Oxford University (Oxford, United Kingdom), while a small proportion (36/313; 11.5%) received the BBV152[19] (commercial name – Covaxin) developed by Bharat Biotech Ltd, India (Table S1A-C). The vaccine identity was unknown for 49 samples. The vaccination dates were unavailable for 12 partially vaccinated and 2 completely vaccinated cases and therefore these 14 samples were excluded during NV analysis of vaccination breakthrough cases

For a pan-India and worldwide analyses of widespread lineages, a dataset comprising of 31546 (India) and 678438 (world) consensus genomes (*fasta* files) for the period March 2021 to October, 2021 were accessed from the publicly available Global Initiative on Sharing All Influenza Data[20] (GISAID, https://www.gisaid.org/) repository; accession IDs for all the sequences submitted to GISAID, from this study are listed in Table S2.

### SARS-CoV-2 RNA extraction and sequencing

Total RNA was isolated in a Biosafety level 2 (BSL-2) environment following standard protocols using the RNA isolation kit (MagRNA-II Viral RNA Extraction Kit, Cat. No. G2M030620; Genes2Me, Gurgaon, India; Molecular RNA Extraction Kit, Cat. No. COVEX 100PS, Q-lineBiotech, India; Nucleic acid extraction kit, Cat. No. A200-96; Zybio Inc.; China) as per manufacturer’s instructions. Each RNA sample was subjected to RT-PCR for multiple viral genes (including E-gene and RNA-dependent RNA polymerase (RDRP) gene) using the nCoV-19 RT-PCR detection kit (Cat. No. NCoV-19ER100PS, Q-lineBiotech, India) or the ViralDetect-II multiplex real time PCR kit for COVID-19 (which also detected the N-gene; Cat. No. G2M020220; Genes2Me, Gurgaon, India). Samples exhibiting a Ct value of <30 (E and RdRp genes) were selected for whole genome sequencing.

The synthesized cDNA was amplified using a multiplex polymerase chain reaction (PCR) protocol, producing 98 amplicons across the SARS-CoV-2 genome (https://artic.network/). The primer pool included additional primers targeting human RNA, producing an additional 11 amplicons. The amplified products were processed for tagmentation and indexing PCR for Illumina Nextera UD Indexes Set A, B, C, D (Illumina Inc., San Diego, California, United States) (384 indexes, 384 samples). All samples were processed as 96-well plate batches that consisted of one each of COVIDSeq positive control HT (CPC HT) and one no template control (NTC); these 96 libraries were pooled, quantified (Qubit 2.0 (Invitrogen, Massachusetts, United States) and fragment sizes were analyzed in Agilent Tapestation 4200 (Agilent Technologies Inc., Santa Clara, California, United States). The pooled library was further normalized to 10nM concentration and 10μl of each normalized pool containing index adapter set A, B, C, and D were combined in a new microcentrifuge tube to a final concentration of 2nM. The pooled libraries were denatured and sequencing was performed on Nextseq 2000 using the P2 100 Cycle kit with 1×101bp Sequencing chemistry. About 50-100Mb data was generated for each sample.

### *In silico* workflow for processing genome sequencing data

The raw sequencing data in *fastq* format was subjected to quality checks including filtering bad quality reads, determination of sequencing depth and adapter trimming using Trimmomatic[21]. All reads shorter than 30 bases or with a Phred quality score <20, were discarded. 18 samples were rejected because of low overall sequencing depth and poor quality. The trimmed reads were then aligned to the reference Wuhan sequence (NCBI ID - NC_045512.2) using bwa-mem[22] algorithm. Post alignment filtering and quality assessment was performed using samtools[23]. Single nucleotide variants (SNVs) were identified using iVar[24] which works on the *mpileup* output from samtools. The variants were annotated using SnpEff[25] and further filtered to remove all problematic sites documented to be prone to accumulate sequencing errors by multiple sources as recommended earlier (https://virological.org/t/issues-with-sars-cov-2-sequencing-data/473). Sequences with high coverage, low N content, and associated with complete metadata were included for further analysis. Reads were assembled to generate consensus *fasta* file using samtools mpileup and the consensus module of iVar with a base assigned as consensus if it had a minimum depth of at least 10 reads (setting ivarMinDepth=10). Sequences where the N content was > 30% and sequence length < 27,000 were rejected. Lineage assignment was done on consensus *fasta* files using Pangolin[26] version (v3.1.16).

All alleles with an allele frequency of >90% were classified as NVs. For analysis of NV cross-correlation, a methodology similar to the one reported in our previous study[14], was used. Briefly, a binary matrix was constructed for each sample with all NVs found in >5% samples as columns, indicating whether an NV of interest was present or absent in a sample. NVs exhibiting low standard deviation across samples were not included in the analysis. Pairwise Pearson correlation coefficients were estimated for this binary matrix using *cor* function in R. p-values indicating significance of association between each pairwise correlation coefficient was estimated using the *cor*.*test* function with *chi-square* test. This matrix was then used to visualize the NV cross-correlation maps in R *corrplot* function[27]. Odds ratio for estimating the association likelihoods of genomic alterations with vaccination breakthrough cases were estimated by creating contingency matrices for each NV identified in >5% of vaccinated samples and were compared with multiple random subsamples of non-vaccinated cases starting March 2021 onwards.

The estimation of intra-host Single Nucleotide Variations (iSNVs) was carried out using Lofreq[28], a variant caller with a high sensitivity to predict iSNVs with an allele frequency as low as 1%[28]. The minor alleles with allele frequencies between 2% and 50%, and minimum sequencing depth of 100x, were classified as iSNVs, and were used for further analysis.

All statistical tests and analyses were performed using custom R scripts. All structural representations were generated in PyMOL (The PyMOL Molecular Graphics System, Schrödinger, LLC).

## Results

### B.1.617.2 (‘Delta’) displaced all previously circulating lineages from March 2021 onwards

We identified a clear shift in the dominant lineages present in Telangana, India, from 2020 to 2021 current year (Figures 1a and S1a; the complete distribution of all lineages in the dataset is provided as supplementary Table S3). The B.1.1.306 and B.1.1.326 lineages (both detected first in India and supposed to have transmission links to Zambia, Somalia and Bahrain (https://cov-lineages.org/lineage_list.html)) were dominant from May, 2020 to September, 2020. The period October, 2020 to March, 2021, witnessed an upsurge of B.1.36.29 (assigned as ‘Indian Lineage’ by Pangolin[26] and pangoLEARN[29]). December, 2020, witnessed the emergence of the Kappa (B.1.617.1) lineage in the state population, and was present till April 2021. However, ‘Alpha’ (B.1.1.7), which was the first lineage classified by WHO as a variant of concern (VoC), appeared in the population in February 2021 and was identified in samples till March 2021, consistent with other reports[30]. Spread of the Alpha lineage was higher and lasted longer in north India, appearing as early as January, 2021, and present till May, 2021, while south India witnessed a higher prevalence of the Kappa variant (B.1.617.1) (Figures S1a,b). However, from March, 2021 onwards, the Delta variant (B.1.617.2) constituted an overwhelming majority of lineages detected in the state, replacing all other virus lineages circulating previously, indicative of its massive spread (Figures 1a and S1a). As expected, the extent of its spread in Telangana almost paralleled its surge in the rest of the country (Figure S1b). Lineage analyses on the two sample sets representing local transmission clusters (Table S1a) did not reveal enrichment of a specific lineage compared to other samples analysed from same period in parallel (data not shown). The spread of Delta in India pre-dated its spread in other countries (Figure 1b). Following its gradual rise in Asia, the Delta lineage spread to Europe, North and South Americas, with the highest spike in June – August, 2021 (Figure 1b). By the end of June, 2021, Delta was the major lineage in all geographical regions of the world[15].

**Figure 1:**
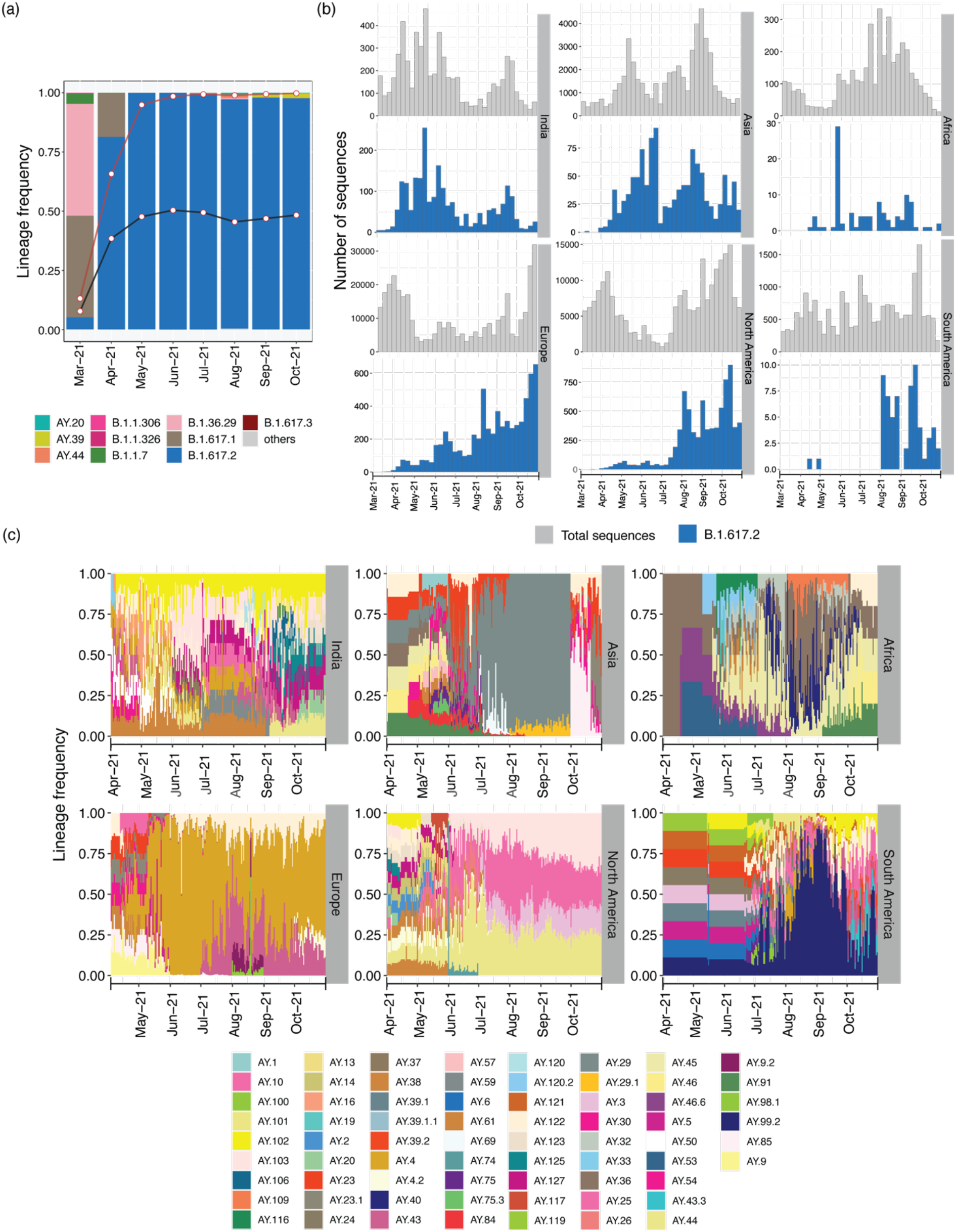
Comparative timeline of major SARS-CoV-2 lineage distribution in Telangana, India. (a) Frequency distribution of the major lineages per month from March, 2021 (a complete timeline starting from April 2020 is provided in Figure S1a). The black and red lines show the frequency distribution trend of the Delta and Delta plus its sub-lineages in India (excluding Telangana), respectively; white points show the corresponding monthly frequency. (b) Timeline of Delta variant distribution (blue) and total (grey) cases in India and rest of the world (all data were obtained from GISAID). (c) Timeline of changing frequency of Delta sub-lineages across the world. Only those sub-lineages present in >5% of total sequences submitted from the region were included. In panels b) and c), while estimating frequencies for the Asian countries, Indian sequences were excluded.

In July 2021, the Delta variant was further classified (by Pangolin) into several sub-lineages (https://cov-lineages.org/) indicative of appearance of additional NVs (https://outbreak.info). From July 2021 onwards, AY.20, AY.39, and AY.44 were the major Delta sub-lineages observed in Telangana (Figure 1a), though the fraction of Delta sub-lineages was higher in most Indian states compared to Telangana (Figure S1b). Moreover, the major sub-lineages in other states varied from July 2021 onwards (Figure S1b).

The distribution of Delta sub-lineages also differed worldwide (Figure 1c). From mid-April, 2021 onwards, AY.4 was most prominent in Europe, followed by AY.43 from July 2021; while AY.103, AY.44, AY.3 and AY.25 were more common in North America from June 2021. The sub-lineage AY.29 was highly prevalent in Oceania from July 2021 (Figure S1c), while Asian countries (excluding India) displayed a mix of various sub-lineages with AY.29 becoming prevalent between June to September, 2021 (Figure 1c). Similar to the distribution pattern observed in India, a mix of different Delta sublineages including AY.36, AY.40, AY.45, AY.46, and AY.91 were observed in sequences from Africa during May-October 2021. Given the consistently increasing repertoire of Delta sub-lineages, constant lineage re-assignment by Pangolin and the fact that a mere assessment of the lineages *per se* may be insufficient to comprehend the complete catalogue of SARS-CoV-2 NVs, we proceeded to perform a detailed analysis of NV profiles obtained from the samples from Telangana, India.

### Genome-wide analysis reveals a sudden change in the landscape SARS-CoV-2 NVs coinciding with the second wave

Study of independent NVs, which may not be included as part of the NV signature of a lineage, and could have arisen due to positive selection, may be critical from the perspective of public health surveillance. A monthly assessment revealed a significant shift in the NVs observed in Telangana during the course of last one year (Figure S2). Overall, a few NVs were consistently present in the population starting from April, 2020, namely A23403G (D614G, S), C241T, C3037T (P924F, ORF1a: nsp3), and C14408T (P4720L, ORF1b: nsp12) (Figure S2). Genes coding for envelope and membrane proteins (E and M genes, respectively) and the accessory protein ORF6, remained relatively free from high frequency NVs till October 2021. The genomic landscape of the virus was marked by the presence of few high frequency NVs in the period from April, 2020 to July, 2020 (Figure S2). The period from August, 2020 to February, 2021 witnessed appearance and subsequent disappearance of many moderately frequent NVs, especially in ORF1ab. However, March, 2021 onwards, a larger number of NVs became evident at moderate to high frequencies throughout the genome (Figure S2). Specifically, the S protein, nucleocapsid (N), ORF7a/b, and nsp3 accumulated several missense NVs (Figure 2). The S protein NVs included T19R, T95I, G142D, del156-157, A222V, L452R, T478K, D614G, P681R, and D950N, along with the ubiquitous D614G (Figure 2). With the exception of T95I, G142D and A222V, all other NVs were consistently present in >75% of samples till October 2021 and were further associated with the appearance of Delta lineage in the population (Figure S3). Beginning May, 2021, additional NVs appeared in the S protein which resulted in bifurcation of Delta into various sub-lineages (Figures 2 and S3). Notably, the frequency of T95I was highest in AY.20 compared to Delta and other sublineages while the frequency of G142D was highest in Kappa lineage (B.1.617.1) compared to other lineages observed thereafter. A222V was another alteration shared by both Delta and AY.44 though it was present in higher frequencies in AY.44 compared to Delta. Interestingly, an observation which clearly stood out from this analysis is that AY.20, AY.39 and AY.44 sub-lineages harbored several NVs (including M153I, A243P, L244F, V1104L, V1176F (in AY.20), S221A, A1080S (in AY.39), G181V (AY.20, AY.39), and T1117I, D1260E (in AY.44)) in the N-terminal domain (NTD), receptor binding domain (RBD) and the region between the two heptapeptide repeat sequences HR1 and HR2, which were either absent or present in lower frequencies in Delta (Figure S3).

**Figure 2:**
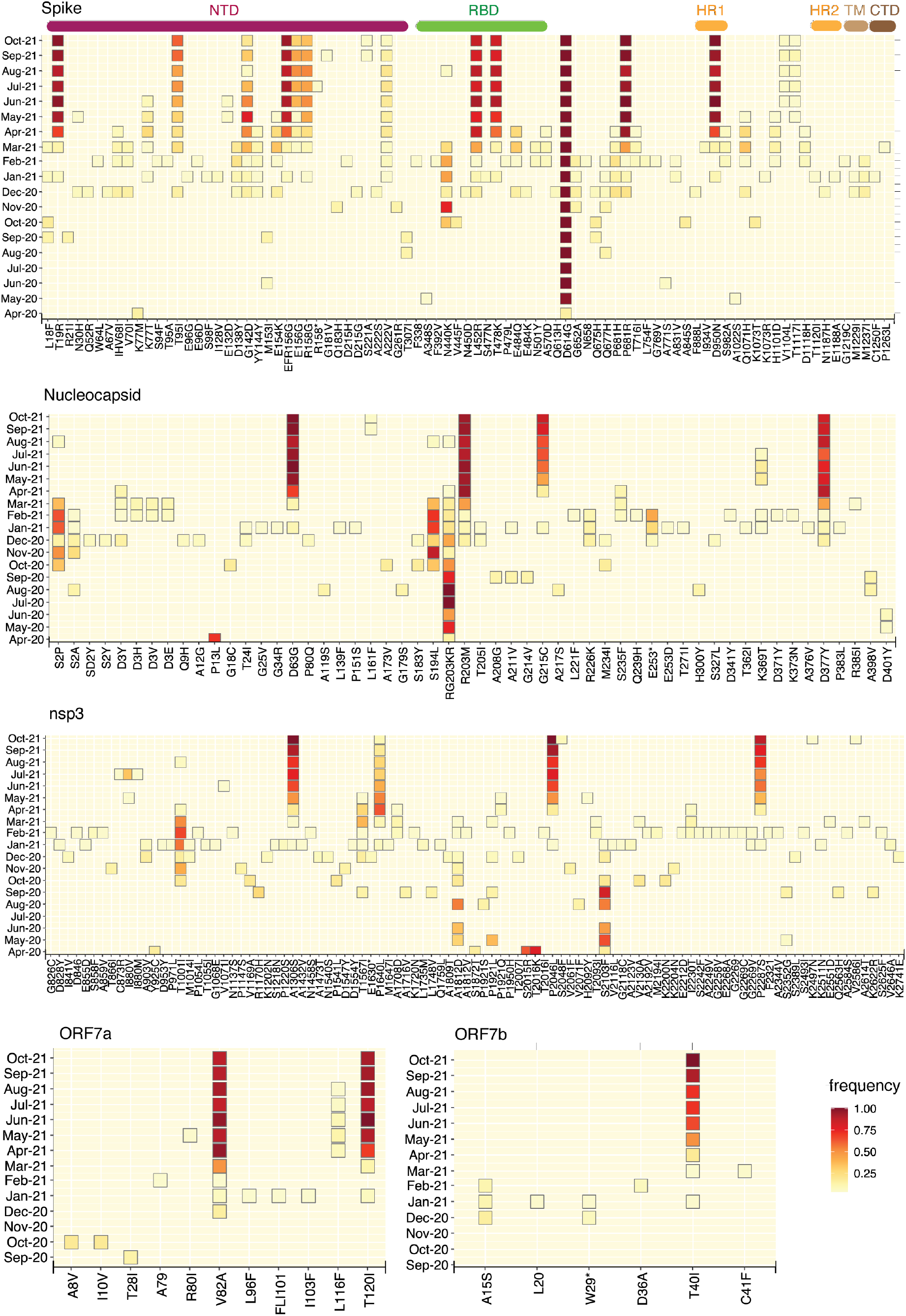
Monthly timeline of SARS-CoV-2 NV frequencies; variants present in <3% of samples were excluded. Key domains in the S protein are indicated at the top: NTD, N-terminal domain (brown); RBD-receptor binding domain (green); S2 subunit containing SD1 (subdomain 1), SD2 (subdomain 2), and S1/S2 cleavage sites (orange); CTD-C-terminal domain (black).

A striking observation in the N protein was the disappearance of RG203KR (triplet 28881-3GGG>AAC) from January, 2021 onwards (Figure 2). We observed the simultaneous appearance of R203M in the Alpha as well as in the Delta and its sub-lineages. Furthermore, the Delta lineage was marked additionally by N protein NVs D63G, G215C, and D377Y. More importantly, several N protein NVs were restricted to Delta sub-lineages (being absent in Delta itself) including L161F, K361Q and K369T (AY.39), and A252S (AY.20) (Figure S3).

Among the non-structural proteins that constitute ORF1a/b, nsp3 is the largest, and exhibited an increase in frequency of several NVs (I880V, A1306S, P1640L, P2046L, P2287S) from March, 2021; most of them being associated with Delta and its sub-lineages (Figures 2 and S3). The accessory proteins 7a and b, relatively free of NVs till December, 2020, accumulated distinct high frequency NVs including V82A (7a), T120I (7a), and T40I (7b) from January 2021 (Figures 2 and S3). Similarly, the NV landscape of ORF3a was significantly altered post-December, 2020 and ORF1b:nsp12 (RNA dependent RNA polymerase) accumulated the G5068S NV in addition to the ubiquitously present P4720L (Figure S4). The frequencies of all these NVs in ORF7a/b, ORF3a, and nsp12, were consistently high in samples classified as either Delta or its sub-lineages AY.20, AY.39, and AY.44, potentially reflecting an extended genomic variation footprint not observed in previous lineages (Figures S3, S4). Furthermore (and similar to the observations made with respect to lineages), an inspection of the NVs in the samples collected from two local ‘super-spreader’ events (Table S1A) did not reveal enrichment of any specific NVs.

In a nutshell, the Delta sub-lineages were marked by presence of several NVs, over and above those that defined the Delta. Secondly, AY.20 and AY.39 had relatively higher numbers of NVs compared to other sub-lineages observed in the state. Since the emergence of vaccine breakthrough cases coincided roughly with emergence of Delta and its sub-lineages, we proceeded to investigate whether the lineages or the NVs associated with them, showed a higher likelihood of occurrence in vaccination breakthrough cases.

### Association of specific genomic NVs with vaccination breakthrough cases

A large fraction (70%) of vaccination breakthrough cases belonged to the Delta lineage, followed by AY.44 (5.2%), AY.20, AY.43, and AY.39 (5% each) (Figure S1d). Interestingly, 57%, 39%, 29%, and 22% of all samples classified as AY.32, AY.43, AY.35, and AY.20 respectively, belonged to vaccination breakthrough cases (Figure S1d). We next analysed all S protein NVs present in >3% of vaccinated cases. T19R, deletion EFR156G (156-157del), L452R, T478K, D614G, P681R, and D950N, present in high frequency in the total dataset, were also present with highest frequencies (>80%) in vaccinated cases (Figure S5), as expected. In addition to these, T95I, which was found to be present in slightly higher frequencies in samples belonging to AY.20 compared to the Delta, was found in 50.8% of all vaccinated samples. Another S protein alteration V1104L (located in the S2 subunit), was present in ∼7.5% of all vaccinated cases. Intriguingly, frequency of this NV was higher in AY.20 associated samples than in Delta itself (Figure S3). A separate analyses of partial and completely vaccinated cases identified S221A (2.3% cases) as an exclusive event in completely vaccinated cases (Figure 3a). Similarly, T1117I (∼3.9% cases) and H1101Y (∼1.9% cases), were identified exclusively in partially vaccinated cases (Figure 3a). We computed odds ratios to estimate significance of association of genome-wide NVs with vaccination breakthrough cases. V1104L (S), S26L (ORF3a), V82A (ORF7a), R203M (N), and T3646A (ORF1ab) exhibited odds ratios ranging between 1.5-3.0 (p-value < 0.05, at 95% confidence interval (Figure 3b); details of upper and lower confidence interval limits are provided in supplementary Table S4), thereby providing further support towards favourable occurrence of these NVs in vaccination breakthrough cases

**Figure 3:**
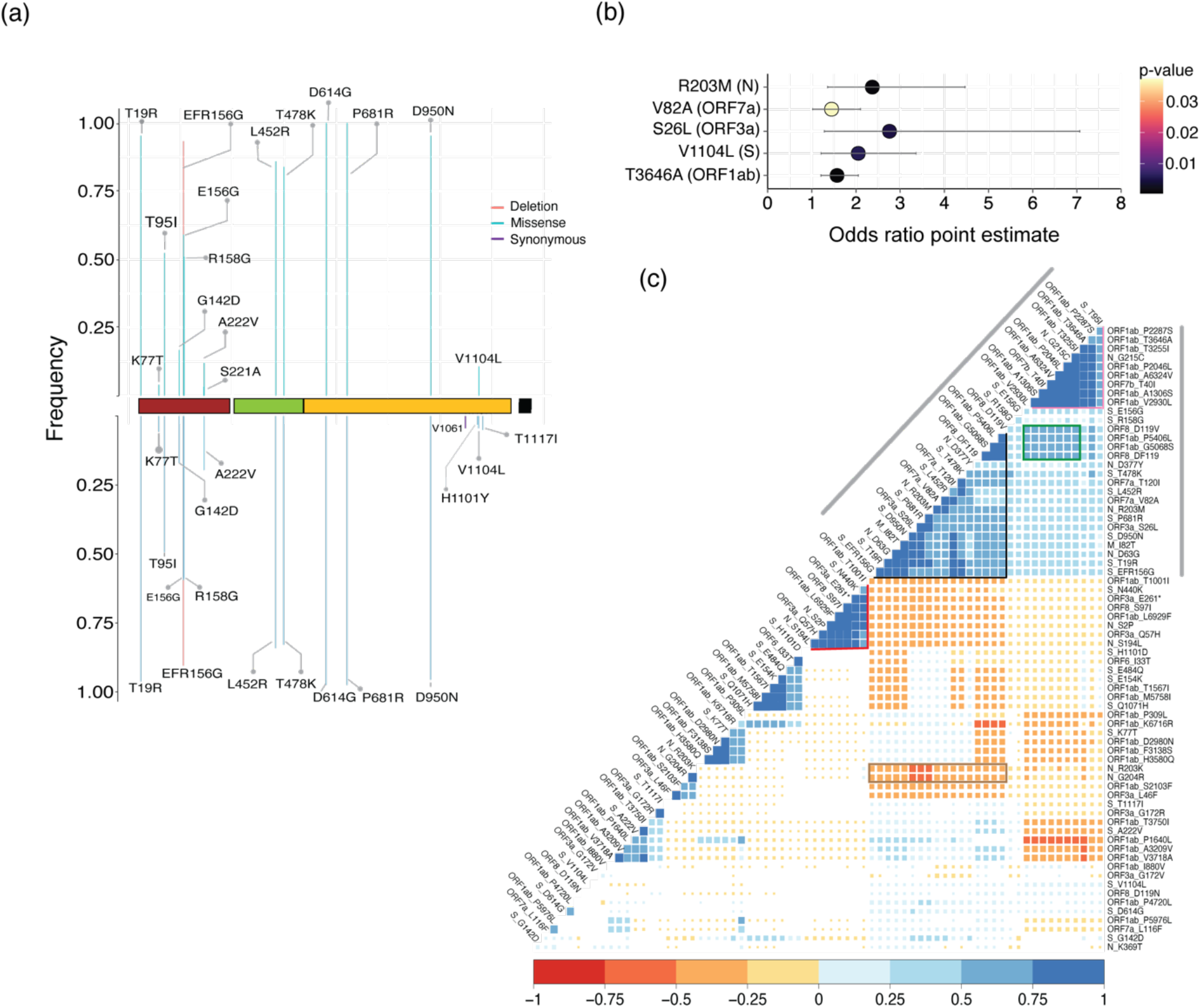
NV frequency in vaccinated cases and pairwise cross-correlation analysis. (a) Frequency of S protein amino acid alterations present in >3% of completely (top) and partially (bottom) vaccinated cases. (b) odds ratio indicating the extent of association of specific variants with vaccination breakthrough cases. (c) Pairwise cross-correlation plot between all non-synonymous missense NVs present in >3% of all the SARS-CoV-2 samples identified from Telangana, India during April 2020 till October, 2021; the size of all coloured ‘squares’ are inversely proportional to the corresponding p-value of the correlation. Positions with p-values > 0.05 appear as blank (or white). The colour key for positive (blue) and negative (red) correlation is given below the plot.

### NV cross-correlation maps reveal presence of extended NV signatures associated with different lineages

An NV (missense only) cross-correlation map arranged by hierarchical clustering revealed several clusters highlighting the frequency of co-occurring NVs within samples. The first major cluster immediately apparent from the cross-correlation map (Figure 3c, grey lines at the sides) was formed by an extensive set of co-occurring NVs that highlight the Delta lineage (https://outbreak.info/situation-reports/delta). Within this large cluster however, we identified presence of two smaller (sub) clusters. One of these (black outline; Figure 3c) encompassed S protein NVs P681R, L452R, T478K, D950N, and T19R which were observed in high frequency in samples belonging to the Delta lineage (Figure S3). This sub-cluster additionally displayed enriched co-occurrence of NVs in other genes namely T120I, V82A (ORF7a), S26L (ORF3a), del 119/120 (ORF8), I82T (M) and D63G and D377Y (N), indicative of a potential extended genomic signature of the Delta lineage. Interestingly, this sub-cluster was mutually exclusive with another set of NVs (N440K (S), S2P, S194L (N), Q57H, stop gained E261* (ORF3a)) that were part of the B.1.36.29 lineage (Figure 3c, the lineage defining NVs highlighted by red outline) which was abundant before the emergence of Delta as mentioned above (Figures 2 and S3). The Delta defining cluster was also negatively correlated to the double amino acid changes in N protein R203K, G204R (brown rectangle; Figure 3c) that was prevalent before the emergence of Delta, indicating a completely unique signature formed by NVs in Delta, mutually exclusive to that of previously circulating viral lineages in the population.

The second sub-cluster (pink outline; Figure 3c) included T95I (S), G215C (N), T40I (ORF7b), and several NVs located in ORF1ab including A1306S, P2046L, P2287S (nsp3), T3255I (nsp4), T3646A (nsp6; also favourably associated with vaccination breakthrough cases (Figure 3b)), and A6324V (nsp14). These NVs were found to be present in high proportion (>90%) of samples associated with Delta sub-lineages AY.20, AY.39, and AY.44 (Figure S3). Though these NVs also exhibited positive correlation with the NVs in the Delta sub-cluster (Figure 3c; black outline) a stronger correlation among the NVs within this new cluster points to a divergent evolution of these sub-lineages in the population. Furthermore, several NVs associated within this Delta sub-lineage sub-cluster displayed positive associations with a set of NVs formed by ORF8 (D119V, del 119/120), and ORF1ab (P5406L, G5068S) (green rectangle; Figure 3c), reflecting additional set of co-occurring common NVs occurring in Delta sub-lineages (AY.20, AY.29, AY.44) (Figure S4).

We extended the analysis to vaccine breakthrough cases and observed a positive cross-correlation between S26L (ORF3a), P4720L (ORF1ab) and D614G (S) (correlation coefficient r^2^ > 0.75) which was absent in non-vaccinated cases (Figure S6). Interestingly, S26L (ORF3a) also exhibited highly significant association with vaccination breakthrough cases (Figure 3b). Another positive cross-correlation was observed among V82A (ORF7a) and S protein NVs, L452R, T478K (r^2^ > 0.75, as opposed to r^2^ < 0.75 in non-vaccinated Delta samples), and T19R, del156-157, T95I (r^2^ > 0.5; < 0.5 in non-vaccinated cases). Finally, another instance of increased positive correlation in vaccination breakthrough cases was formed by R203M (N), V82A, T120I (ORF7a) compared to non-vaccinated cases (Figure S6). This aligns well with our previous observation, where both R203M and V82A display favourable odds of occurrence in vaccination breakthrough cases (Figure 3b). Overall, the findings suggest that the genomic alterations in S protein (L452R, T478K, P681R) which have previously been reported to increase escape from neutralizing antibodies and potentially associate with vaccination breakthroughs[7],[10] might arise in tandem with genomic changes located in non-S genes which are involved in modulating the downstream processes in viral life cycle (ORF1ab, N) and regulating host immune response (ORF7a/b, ORF3a), thus increasing the probability of virus survival and causing breakthroughs.

### SARS-CoV-2 exhibits genomic plasticity as multiple sites contribute to generation of intra-host variants

Since virus genomic diversity arises within a host, it becomes important to uncover minor alleles originating in the form of iSNVs. A total of 545 samples (15.4% of the total dataset) exhibited iSNVs of which a majority (234; 43% (6.6% of total dataset)) harboured minor allelic variants at a single genomic position and 165 samples (30% (4.6% of the total dataset)) exhibited iSNVs at two positions (Figure 4a). Interestingly, 11 samples contained more than 9 sites with minor alleles. However, the Ct values in 7 of these 11 cases were > 22 (Table S5) indicating possible false calls due to sequencing error(s), as reported earlier[31–33]. Presence of >9 iSNVs sites in rest of the 3 samples could probably reflect mixed infections, as reported earlier[31]. N, nsp11, nsp9, and ORF3a exhibited higher frequency of iSNVs (when normalized to gene size) compared to other proteins (Table S6).

**Figure 4:**
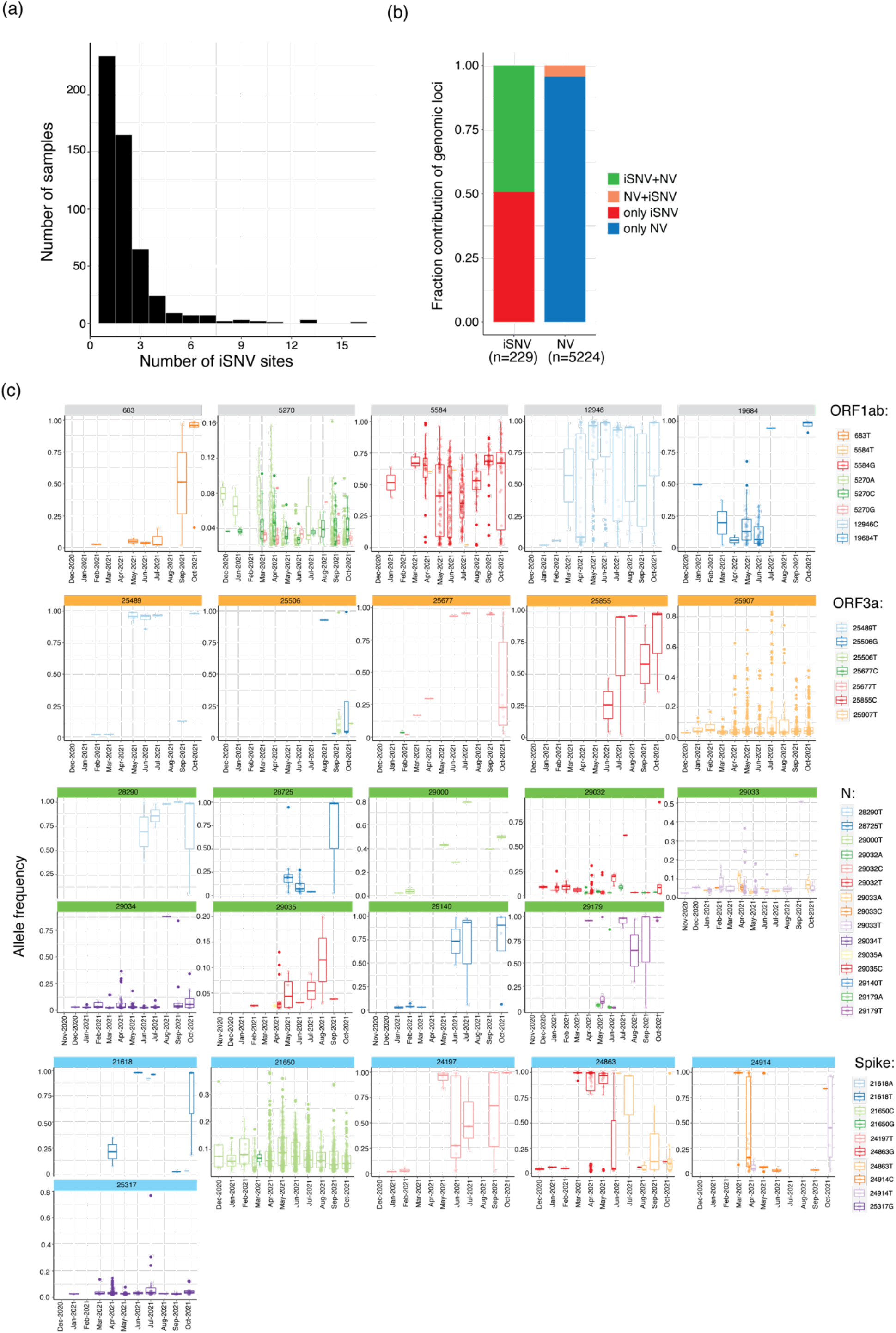
SARS-CoV-2 iSNVs identified from Telangana, India. (a) Distribution of number of iSNVs in samples. (b) Distribution of genomic loci with shared and unique iSNVs and NVs. (c) Timeline of allele frequency changes in the minor alleles in ORF1ab, ORF3a, N, and S proteins. Box plots indicate allelic frequency distribution while the points represent samples in which the allele was identified. Only those alternate alleles whose allele frequencies were either consistent or increased during the indicated timeline are shown.

We next endeavoured to map co-occurrence of NVs and iSNVs in the genome. Across the 545 samples, a total of 229 genomic loci were involved in iSNVs, of which 113 loci were found to also share NVs (Figure 4b). This observation stands in contrast to total number of genomic loci which formed NVs across the dataset (5224), of which only 229 sites shared iSNVs. In order to investigate whether any iSNV site(s) which shared NVs coincided with mutation ‘hotspot’ region(s), we mapped all genomic positions which shared iSNV and NVs and estimated the sample frequencies of major and alternative alleles at these loci (Figure S7). The analysis revealed that a very few (9053 (nsp4), 11201, 11332, 11418 (nsp6), 21618, 23403, 23604 (S), 26767 (N), and 28881 (N)) shared sites exhibited NVs (major alleles) which were widespread (sample frequency > 45%) in the dataset suggesting that these loci could be mutation hotspots.

Another critical, and arguably more important insight iSNV analysis provides is by enabling an effective temporal tracking of a novel minor allele from the time of its emergence. This can potentially identify an important NV before it becomes widespread in the population; hence being of great benefit for public health surveillance. To this end, we evaluated changes in sample and allele frequencies of minor alleles identified in iSNV sites (Figure 4c). Results revealed few iSNVs in ORF1ab (683T, 5584G, 12946C, and 19684T), ORF3a (25489T, 25506T, 25506G, 25677T, 25855C, and 25907T), and multiple positions in N (28290T, 28725T, 29000T, 29032T, 29034T, 29140T, and 29179T) where the sample and allele frequencies consistently increased from the time of their emergence (Figure 4c). Allele frequencies of minor alleles at all these iSNV positions (with a few exceptions discussed below) reached 80% over time attesting to a transition from iSNV to NV. Furthermore, most of these iSNVs emerged in January 2021 (with few emerging in December 2020 or February 2021), indicating towards a possible escape from transmission bottleneck. Few notable examples include 25907T (leading to 172V in ORF3a, present in 53% of total samples) and 29034T in N (leading to 254V, present in 24% of all samples).

In addition, a few iSNVs exhibited a significant increase in sample frequency across a long time period, despite exhibiting an allele frequency of < 50% (Figure 4c). A notable example being all alternative alleles at position 5270 (nsp3), which exhibited allele frequency <16%, but were nonetheless detected in a significant fraction of samples (across the 3543 samples, 5270A in 33% cases, 5270C in 20%, 5270G in 5%) (Figure 4c). Another such instance was observed at position 29033 (N; 29033T in 6%, 29033A in 1.2%) (Figure 4c).

We next focused on S protein iSNVs, expected to be important in shaping virus transmission and immune escape. In addition, S protein iSNVs are reported to be rare events due to evolutionary constraints with many getting lost attesting to a narrow transmission bottleneck[34]. A few minor alleles 19I (C21618), 30H (A21650), 879S (G24197), 1101Y (C24863), 1118Y (C24914), and 1252C (C25317), which were first observed in independent samples in January 2021, were consistently present in samples till October 2021 as well (sample distribution and allele frequency shown in Figure 4c). Among these, the S protein iSNVs associated minor alleles that exhibited an increased sample frequency from the time of their emergence were 30H, 1101Y, and 1252C; the distribution of their allelic frequencies in shown in Figure 4c. However, the allelic frequencies of 30H and 1252C, in all the samples in which they were detected, was < 50% (with the exception of one sample which harbored the 1252C allele at allele frequency > 75%). On the contrary, from June to September 2021, the sample frequency of 1101Y increased from 0.6% to 3%, with a concordant increase in corresponding allele frequency (Figure 4c). Interestingly, the H1101Y NV was also present in >5% of all partially vaccinated samples. Subsequent analysis on the potential functional impact of these iSNVs is currently underway.

## Discussion

SARS-CoV-2 continues to cause significant morbidity and mortality worldwide making it important to perform regular genomic surveillance to detect emerging virus NVs. The estimated mutation rate of SARS-CoV-2 is about 1.1 × 10^−3^ substitutions/site/year[35]. The constant virus evolution leading to emergence of new variants is shaped by several factors including host genetics and immune response[36] [37], strong selection pressure created due to neutralizing antibodies[38], etc. The infection rate reduced significantly across India after peaking in August, 2020. However, India witnessed a ferocious ‘second wave’ of infection during February to June, 2021, with number of deaths several folds higher than that observed in the first wave[39]. The massive spread has mostly been attributed to the Delta variant, which has been shown to be associated with higher transmissibility rates than previously circulating Alpha and Kappa lineages[40], [41]. The Delta rapidly displaced all previously circulating viral lineages owing perhaps to its increased fitness (R0 (Re) 60-70%) with viremia 1000 folds higher than most previous lineages[41]. The increased adaptability of a pathogen virus under ‘waning’ immune pressure or partial immunization, has been reported in other studies[42]. While other widely transmitted lineages such as the Alpha emerged in other countries before vaccination programs were implemented[5], the countrywide spread of the Delta probably occurred during the implementation of vaccination in India. This suggests that higher mutation accumulation observed in this variant could be a result of increased selection pressure under a modified host microenvironment which the virus was exposed to. This suggestion assumes significance given the Delta variant’s ultrafast replication speeds which makes it detectable within 4 days after exposure[41]. Despite the preponderance of Delta during the second wave, the spread of specific virus lineage(s)/sub-lineages did exhibit differences in various geographical regions within India. Under these circumstances, it becomes imperative to study the evolution of the virus within a specific demography over an extended time period.

From June 2021 onwards, the Delta variant itself was divided into several sub-lineages (based on Pangolin classification) but we have presented results mainly for the more frequently observed ones namely AY.20, AY.39, and AY.44. The preferential occurrence of certain sub-lineages over others in different states within India and in different countries indicates a potential role of population-specific host genetic factors which might govern the favourable spread of one sub-lineage over another. We have however not evaluated the association between host genetics and viral sub-lineages in this study. Although there are multiple reports of the association of a few S protein NVs (especially T478K, P681R, and L452R) with viral transmissibility and immune escape[10],[43], we have performed an extensive analyses on the entire landscape of SARS-CoV-2 genomic NVs in this study. Although these NVs have been reported to occur in samples belonging to Delta lineage (as documented in GISAID, and https://outbreak.info/situation-reports/delta), their association with vaccination breakthrough events were not reported thus signifying the importance of our analysis. However, absence of information on neutralization antibody levels in vaccinated individuals did make it difficult for us to establish a strong association with vaccination breakthrough cases. Also, keeping in mind the interpretation pitfalls that may be created due to the small size of vaccination breakthrough cases, we are currently validating the results presented here on a larger sample set.

The NV cross-correlation analysis is a powerful tool to establish moderately or tightly linked genome-wide signatures. Our analyses revealed the complete footprint of genomic alterations affiliated to specific viral lineages. The cross-correlation analysis not only described the entire set of alterations associated with Delta and its sub-lineages, but also revealed several NVs that were completely mutually exclusive with those associated with previously circulating lineages. Another significant observation made possible through the cross-correlation analysis was the preferential co-occurrence of specific NVs in Delta sub-lineages but absent from Delta itself; which could not be deduced from a linear analysis of NV timeline. The analyses also suggested higher enrichment of certain co-occurring NVs in vaccination breakthrough (compared to other) cases.

Previous studies have shed evidence on how iSNVs impart genomic plasticity[44] and direct viral genome evolution through inter-host transmission cycles[45,46]. Due to lack of data on donor-recipient pairs and primary contacts (including family members) of infected individuals, we were unable to perform iSNV bottleneck estimation in this study. Moreover, despite having a substantial dataset size, the information output is hampered by lack of clinical information like infection symptoms, hospitalization status, etc. Nevertheless, few observations are worth highlighting. Firstly, we did not detect significant correlation between samples showing iSNVs and their vaccination status or their age or with a specific lineage (data not shown). Secondly, we identified specific iSNVs like the 25907T in ORF3a that exhibited increased sample and allele frequencies with time. Interestingly, the 172V alteration generated from 25907T has been shown to improve protein stability, owing to increased local hydrophobic interactions, in recent studies[47]. The stability of ORF3a, plays a crucial role in its functionality as an apoptosis inducing protein leading to cell death[48] and membrane rearrangement during SARS-CoV-2 infection[49].

The iSNV analysis has potentially revealed an important S protein allele viz. 1101Y, where the ‘Y’ allele frequency showed an upward trend from April 2021 and was labelled as an NV as its frequency became >50%. Further, this NV was also preferentially associated with partially vaccinated samples. Interestingly, H1101Y, V1104L, and T1117I are located between the two heptapeptide repeat sequences HR1 and HR2 within the S2 subunit of the S protein (Figure S8). Earlier reports have suggested that both V1104L and H1101Y could increase local stability and alter the surface character of the S protein, thereby aiding in favourable evolution of the virus[50,51]. We therefore recommend to include H1101Y and V1104L under active surveillance.

This study provides fresh insights into how the virus genome landscape has evolved over the duration of 19 months in a localized population in India. The novelty of the study stems from its all all-inclusive approach and the identification of missense NVs in the context of cross-correlation and intra-host diversity analysis. More importantly, we laid greater emphasis on individual NVs rather than the lineages per se; given the recent and frequent ‘re-classifications’ of SARS-CoV-2 lineages by Pangolin[29]. Our study has facilitated better understanding of how different aspects of virus genome dynamics are inter-linked. Future functional studies on important NVs identified in this study may reveal their possible role(s) in virus transmission and vaccine escape. Given the ongoing immunization program, it may be worthwhile to perform similar studies in other regions of India.

## Data Availability

All data produced in the present work are available in the Sequence Read Archive (SRA) bioproject no - PRJNA691556

## Acknowledgements

Authors acknowledge all patients who consented to be part of this study. We are grateful to Dr Nagamani, Telangana state nodal officer, for coordinating sample collection from across the state. We thank all Telangana state sentinel sites assigned for identifying samples for genome sequencing namely Gandhi Medical College and Hospital, Secunderabad; Institute of Preventive Medicine, Hyderabad, Osmania Medical College, Hyderabad, Nizam’s Institute of Medical Sciences (NIMS), Hyderabad, Fever Hospital, Hyderabad, Government Medical College (GMC), Siddipet, Government Medical College, Warangal, Government General Hospital and Medical College, Suryapet, Government Medical College (GMC), Mahbubnagar, Government Medical College, Nizamabad. We also acknowledge Dr S. Raju (State Public Health Laboratory (SPHL, Chennai, TN) and Dr Gyaneshwar Chaubey, Banaras Hindu University (BHU, UP), for access to vaccination breakthrough samples. We thank the Telangana State Government, the Indian Council of Medical Research, Government of India, and the University of Hyderabad, Hyderabad, for procurement of consumables and equipment to perform screening of patient samples. We are grateful to Dr K Thangaraj, Dr Ashwin B Dalal, Dr Rashna Bhandari and Dr R Harinarayanan, CDFD, Hyderabad, for co-ordinating the activities of the COVID-19 testing laboratory at CDFD. We are grateful to Mr Mandla Vasanth Kumar for his kind assistance in computational analysis. All volunteers and ‘COVID warriors’ from CDFD, Hyderabad, are gratefully acknowledge for their significant contribution in screening of samples. We also acknowledge the National Genomics Core (NGC) – CDFD for performing SARS-CoV-2 genome sequencing.

## Author Contributions

M.D.B. and A.G. conceptualized the study. A.G., M.D.B., and R.B. developed the methodology. A.G. carried out the formal analysis. Writing (Original draft preparation) was carried out by A.G.. M.D.B., and A.G. worked on reviewing and edited the draft, R.B. provided suggestions on editing the draft.

## Data availability

All the raw sequencing data used in this study have been submitted to the Sequencing Read Archive (SRA) with project accession ID PRJNA691556.

## References

1 Frampton D, Rampling T, Cross A, Bailey H, Heaney J, Byott M, Scott R, Sconza R, Price J, Margaritis M, Bergstrom M, Spyer MJ, Miralhes PB, Grant P, Kirk S, Valerio C, Mangera Z, Prabhahar T, Moreno-Cuesta J, Arulkumaran N, Singer M, Shin GY, Sanchez E, Paraskevopoulou SM, Pillay D, McKendry RA, Mirfenderesky M, Houlihan CF & Nastouli E (2021) Genomic characteristics and clinical effect of the emergent SARS-CoV-2 B.1.1.7 lineage in London, UK: a whole-genome sequencing and hospital-based cohort study. The Lancet Infectious Diseases, S1473309921001705.

2 Cherian S, Potdar V, Jadhav S, Yadav P, Gupta N, Das M, Rakshit P, Singh S, Abraham P, Panda S, & NIC team (2021) Convergent evolution of SARS-CoV-2 spike mutations, L452R, E484Q and P681R, in the second wave of COVID-19 in Maharashtra, India Molecular Biology.

3 Davies NG, Abbott S, Barnard RC, Jarvis CI, Kucharski AJ, Munday JD, Pearson CAB, Russell TW, Tully DC, Washburne AD, Wenseleers T, Gimma A, Waites W, Wong KLM, van Zandvoort K, Silverman JD, CMMID COVID-19 Working Group1‡, COVID-19 Genomics UK (COG-UK) Consortium‡, Diaz-Ordaz K, Keogh R, Eggo RM, Funk S, Jit M, Atkins KE & Edmunds WJ (2021) Estimated transmissibility and impact of SARS-CoV-2 lineage B.1.1.7 in England. Science 372, eabg3055.

4 Kumar V, Singh J, Hasnain SE & Sundar D (2021) Possible link between higher transmissibility of B.1.617 and B.1.1.7 variants of SARS-CoV-2 and increased structural stability of its spike protein and hACE2 affinity Biophysics.

5 The COVID-19 Genomics UK (COG-UK) consortium, Volz E, Mishra S, Chand M, Barrett JC, Johnson R, Geidelberg L, Hinsley WR, Laydon DJ, Dabrera G, O’Toole Á, Amato R, Ragonnet-Cronin M, Harrison I, Jackson B, Ariani CV, Boyd O, Loman NJ, McCrone JT, Gonçalves S, Jorgensen D, Myers R, Hill V, Jackson DK, Gaythorpe K, Groves N, Sillitoe J, Kwiatkowski DP, Flaxman S, Ratmann O, Bhatt S, Hopkins S, Gandy A, Rambaut A & Ferguson NM (2021) Assessing transmissibility of SARS-CoV-2 lineage B.1.1.7 in England. Nature 593, 266–269.

6 Torjesen I (2021) Covid-19: Delta variant is now UK’s most dominant strain and spreading through schools. BMJ, n1445.

7 Weisblum Y, Schmidt F, Zhang F, DaSilva J, Poston D, Lorenzi JC, Muecksch F, Rutkowska M, Hoffmann H-H, Michailidis E, Gaebler C, Agudelo M, Cho A, Wang Z, Gazumyan A, Cipolla M, Luchsinger L, Hillyer CD, Caskey M, Robbiani DF, Rice CM, Nussenzweig MC, Hatziioannou T & Bieniasz PD (2020) Escape from neutralizing antibodies by SARS-CoV-2 spike protein variants. eLife 9, e61312.

8 Xie X, Zou J, Fontes-Garfias CR, Xia H, Swanson KA, Cutler M, Cooper D, Menachery VD, Weaver S, Dormitzer PR & Shi P-Y (2021) Neutralization of N501Y mutant SARS-CoV-2 by BNT162b2 vaccine-elicited sera Microbiology.

9 Zhou D, Dejnirattisai W, Supasa P, Liu C, Mentzer AJ, Ginn HM, Zhao Y, Duyvesteyn HME, Tuekprakhon A, Nutalai R, Wang B, Paesen GC, Lopez-Camacho C, Slon-Campos J, Hallis B, Coombes N, Bewley K, Charlton S, Walter TS, Skelly D, Lumley SF, Dold C, Levin R, Dong T, Pollard AJ, Knight JC, Crook D, Lambe T, Clutterbuck E, Bibi S, Flaxman A, Bittaye M, Belij-Rammerstorfer S, Gilbert S, James W, Carroll MW, Klenerman P, Barnes E, Dunachie SJ, Fry EE, Mongkolsapaya J, Ren J, Stuart DI & Screaton GR (2021) Evidence of escape of SARS-CoV-2 variant B.1.351 from natural and vaccine-induced sera. Cell 184, 2348–2361.e6.

10 Planas D, Veyer D, Baidaliuk A, Staropoli I, Guivel-Benhassine F, Rajah MM, Planchais C, Porrot F, Robillard N, Puech J, Prot M, Gallais F, Gantner P, Velay A, Le Guen J, Kassis-Chikhani N, Edriss D, Belec L, Seve A, Courtellemont L, Péré H, Hocqueloux L, Fafi-Kremer S, Prazuck T, Mouquet H, Bruel T, Simon-Lorière E, Rey FA & Schwartz O (2021) Reduced sensitivity of SARS-CoV-2 variant Delta to antibody neutralization. Nature 596, 276–280.

11 Edara V-V, Pinsky BA, Suthar MS, Lai L, Davis-Gardner ME, Floyd K, Flowers MW, Wrammert J, Hussaini L, Ciric CR, Bechnak S, Stephens K, Graham BS, Bayat Mokhtari E, Mudvari P, Boritz E, Creanga A, Pegu A, Derrien-Colemyn A, Henry AR, Gagne M, Douek DC, Sahoo MK, Sibai M, Solis D, Webby RJ, Jeevan T & Fabrizio TP (2021) Infection and Vaccine-Induced Neutralizing-Antibody Responses to the SARS-CoV-2 B.1.617 Variants. N Engl J Med 385, 664–666.

12 Cele S, Jackson L, Khoury DS, Khan K, Moyo-Gwete T, Tegally H, San JE, Cromer D, Scheepers C, Amoako D, Karim F, Bernstein M, Lustig G, Archary D, Smith M, Ganga Y, Jule Z, Reedoy K, Hwa S-H, Giandhari J, Blackburn JM, Gosnell BI, Karim SSA, Hanekom W, NGS-SA, COMMIT-KZN Team, von Gottberg A, Bhiman J, Lessells RJ, Moosa M-YS, Davenport MP, de Oliveira T, Moore PL & Sigal A (2021) SARS-CoV-2 Omicron has extensive but incomplete escape of Pfizer BNT162b2 elicited neutralization and requires ACE2 for infection Infectious Diseases (except HIV/AIDS).

13 Kirola L (2021) Genetic emergence of B.1.617.2 in COVID-19. New Microbes and New Infections 43, 100929.

14 Gupta A, Sabarinathan R, Bala P, Donipadi V, Vashisht D, Katika MR, Kandakatla M, Mitra D, Dalal A & Bashyam MD (2021) A comprehensive profile of genomic variations in the SARS-CoV-2 isolates from the state of Telangana, India. Journal of General Virology 102.

15 Mlcochova P, Kemp S, Dhar MS, Papa G, Meng B, Ferreira IATM, Datir R, Collier DA, Albecka A, Singh S, Pandey R, Brown J, Zhou J, Goonawardane N, Mishra S, Whittaker C, Mellan T, Marwal R, Datta M, Sengupta S, Ponnusamy K, Radhakrishnan VS, Abdullahi A, Charles O, Chattopadhyay P, Devi P, Caputo D, Peacock T, Wattal DC, Goel N, Satwik A, Vaishya R, Agarwal M, The Indian SARS-CoV-2 Genomics Consortium (INSACOG), Chauhan H, Dikid T, Gogia H, Lall H, Verma K, Dhar MS, Singh MK, Soni N, Meena N, Madan P, Singh P, Sharma R, Sharma R, Kabra S, Kumar S, Kumari S, Sharma U, Chaudhary U, Sivasubbu S, Scaria V, Wattal C, Oberoi JK, Raveendran R, Datta S, Das S, Maitra A, Chinnaswamy S, Biswas NK, Parida A, Raghav SK, Prasad P, Sarin A, Mayor S, Ramakrishnan U, Palakodeti D, Seshasayee ASN, Thangaraj K, Bashyam MD, Dalal A, Bhat M, Shouche Y, Pillai A, Abraham P, Atul PV, Cherian SS, Desai AS, Pattabiraman C, Manjunatha MV, Mani RS, Udupi GA, Nandicoori V, Bharadwaj K, Tallapaka Sowpati DT, The Genotype to Phenotype Japan (G2P-Japan) Consortium, Kawabata R, Morizako N, Sadamasu K, Asakura H, Nagashima M, Yoshimura K, Ito J, Kimura I, Uriu K, Kosugi Y, Suganami M, Oide A, Yokoyama M, Chiba M, Saito A, Butlertanaka EP, Tanaka YL, Ikeda T, Motozono C, Nasser H, Shimizu R, Yuan Y, Kitazato K, Hasebe H, Nakagawa S, Wu J, Takahashi M, Fukuhara T, Shimizu K, Tsushima K, Kubo H, Shirakawa K, Kazuma Y, Nomura R, Horisawa Y, Takaori-Kondo A, Tokunaga K, Ozono S, The CITIID-NIHR BioResource COVID-19 Collaboration, Baker S, Dougan G, Hess C, Kingston N, Lehner PJ, Lyons PA, Matheson NJ, Owehand WH, Saunders C, Summers C, Thaventhiran JED, Toshner M, Weekes MP, Maxwell P, Shaw A, Bucke A, Calder J, Canna L, Domingo J, Elmer A, Fuller S, Harris J, Hewitt S, Kennet J, Jose S, Kourampa J, Meadows A, O’Brien C, Price J, Publico C, Rastall R, Ribeiro C, Rowlands J, Ruffolo V, Tordesillas H, Bullman B, Dunmore BJ, Fawke S, Gräf S, Hodgson J, Huang C, Hunter K, Jones E, Legchenko E, Matara C, Martin J, Mescia F, O’Donnell C, Pointon L, Pond N, Shih J, Sutcliffe R, Tilly T, Treacy C, Tong Z, Wood J, Wylot M, Bergamaschi L, Betancourt A, Bower G, Cossetti C, De Sa A, Epping M, Fawke S, Gleadall N, Grenfell R, Hinch A, Huhn O, Jackson S, Jarvis I, Krishna B, Lewis D, Marsden J, Nice F, Okecha G, Omarjee O, Perera M, Potts M, Richoz N, Romashova V, Yarkoni NS, Sharma R, Stefanucci L, Stephens J, Strezlecki M, Turner L, De Bie EMDD, Bunclark K, Josipovic M, Mackay M, Michael A, Rossi S, Selvan M, Spencer S, Yong C, Allison J, Butcher H, Caputo D, Clapham-Riley D, Dewhurst E, Furlong A, Graves B, Gray J, Ivers T, Kasanicki M, Le Gresley E, Linger R, Meloy S, Muldoon F, Ovington N, Papadia S, Phelan I, Stark H, Stirrups KE, Townsend P, Walker N, Webster J, Scholtes I, Hein S, King R, Mavousian A, Lee JH, Bassi J, Silacci-Fegni C, Saliba C, Pinto D, Irie T, Yoshida I, Hamilton WL, Sato K, Bhatt S, Flaxman S, James LC, Corti D, Piccoli L, Barclay WS, Rakshit P, Agrawal A & Gupta RK (2021) SARS-CoV-2 B.1.617.2 Delta variant replication and immune evasion. Nature.

16 Pouwels KB, Pritchard E, Matthews PC, Stoesser N, Eyre DW, Vihta K-D, House T, Hay J, Bell JI, Newton JN, Farrar J, Crook D, Cook D, Rourke E, Studley R, Peto TEA, Diamond I & Walker AS (2021) Effect of Delta variant on viral burden and vaccine effectiveness against new SARS-CoV-2 infections in the UK. Nat Med.

17 Nordström P, Ballin M & Nordström A (2021) Effectiveness of heterologous ChAdOx1 nCoV-19 and mRNA prime-boost vaccination against symptomatic Covid-19 infection in Sweden: A nationwide cohort study. The Lancet Regional Health - Europe, 100249.

18 Voysey M, Clemens SAC, Madhi SA, Weckx LY, Folegatti PM, Aley PK, Angus B, Baillie VL, Barnabas SL, Bhorat QE, Bibi S, Briner C, Cicconi P, Collins AM, Colin-Jones R, Cutland CL, Darton TC, Dheda K, Duncan CJA, Emary KRW, Ewer KJ, Fairlie L, Faust SN, Feng S, Ferreira DM, Finn A, Goodman AL, Green CM, Green CA, Heath PT, Hill C, Hill H, Hirsch I, Hodgson SHC, Izu A, Jackson S, Jenkin D, Joe CCD, Kerridge S, Koen A, Kwatra G, Lazarus R, Lawrie AM, Lelliott A, Libri V, Lillie PJ, Mallory R, Mendes AVA, Milan EP, Minassian AM, McGregor A, Morrison H, Mujadidi YF, Nana A, O’Reilly PJ, Padayachee SD, Pittella A, Plested E, Pollock KM, Ramasamy MN, Rhead S, Schwarzbold AV, Singh N, Smith A, Song R, Snape MD, Sprinz E, Sutherland RK, Tarrant R, Thomson EC, Török ME, Toshner M, Turner DPJ, Vekemans J, Villafana TL, Watson MEE, Williams CJ, Douglas AD, Hill AVS, Lambe T, Gilbert SC, Pollard AJ, Aban M, Abayomi F, Abeyskera K, Aboagye J, Adam M, Adams K, Adamson J, Adelaja YA, Adewetan G, Adlou S, Ahmed K, Akhalwaya Y, Akhalwaya S, Alcock A, Ali A, Allen ER, Allen L, Almeida Tcdsc, Alves MPS, Amorim F, Andritsou F, Anslow R, Appleby M, Arbe-Barnes EH, Ariaans MP, Arns B, Arruda L, Azi P, Azi L, Babbage G, Bailey C, Baker KF, Baker M, Baker N, Baker P, Baldwin L, Baleanu I, Bandeira D, Bara A, Barbosa MAS, Barker D, Barlow GD, Barnes E, Barr AS, Barrett JR, Barrett J, Bates L, Batten A, Beadon K, Beales E, Beckley R, Belij-Rammerstorfer S, Bell J, Bellamy D, Bellei N, Belton S, Berg A, Bermejo L, Berrie E, Berry L, Berzenyi D, Beveridge A, Bewley KR, Bexhell H, Bhikha S, Bhorat AE, Bhorat ZE, Bijker E, Birch G, Birch S, Bird A, Bird O, Bisnauthsing K, Bittaye M, Blackstone K, Blackwell L, Bletchly H, Blundell CL, Blundell SR, Bodalia P, Boettger BC, Bolam E, Boland E, Bormans D, Borthwick N, Bowring F, Boyd A, Bradley P, Brenner T, Brown P, Brown C, Brown-O’Sullivan C, Bruce S, Brunt E, Buchan R, Budd W, Bulbulia YA, Bull M, Burbage J, Burhan H, Burn A, Buttigieg KR, Byard N, Cabera Puig I, Calderon G, Calvert A, Camara S, Cao M, Cappuccini F, Cardoso JR, Carr M, Carroll MW, Carson-Stevens A, Carvalho Y de M, Carvalho JAM, Casey HR, Cashen P, Castro T, Castro LC, Cathie K, Cavey A, Cerbino-Neto J, Chadwick J, Chapman D, Charlton S, Chelysheva I, Chester O, Chita S, Cho J-S, Cifuentes L, Clark E, Clark M, Clarke A, Clutterbuck EA, Collins SLK, Conlon CP, Connarty S, Coombes N, Cooper C, Cooper R, Cornelissen L, Corrah T, Cosgrove C, Cox T, Crocker WEM, Crosbie S, Cullen L, Cullen D, Cunha Drmf, Cunningham C, Cuthbertson FC, Da Guarda Snf, da Silva LP, Damratoski BE, Danos Z, Dantas MTDC, Darroch P, Datoo MS, Datta C, Davids M, Davies SL, Davies H, Davis E, Davis J, Davis J, De Nobrega Mmd, De Oliveira Kalid LM, Dearlove D, Demissie T, Desai A, Di Marco S, Di Maso C, Dinelli MIS, Dinesh T, Docksey C, Dold C, Dong T, Donnellan FR, Dos Santos T, dos Santos TG, Dos Santos EP, Douglas N, Downing C, Drake J, Drake-Brockman R, Driver K, Drury R, Dunachie SJ, Durham BS, Dutra L, Easom NJW, van Eck S, Edwards M, Edwards NJ, El Muhanna OM, Elias SC, Elmore M, English M, Esmail A, Essack YM, Farmer E, Farooq M, Farrar M, Farrugia L, Faulkner B, Fedosyuk S, Felle S, Feng S, Ferreira Da Silva C, Field S, Fisher R, Flaxman A, Fletcher J, Fofie H, Fok H, Ford KJ, Fowler J, Fraiman PHA, Francis E, Franco MM, Frater J, Freire MSM, Fry SH, Fudge S, Furze J, Fuskova M, Galian-Rubio P, Galiza E, Garlant H, Gavrila M, Geddes A, Gibbons KA, Gilbride C, Gill H, Glynn S, Godwin K, Gokani K, Goldoni UC, Goncalves M, Gonzalez IGS, Goodwin J, Goondiwala A, Gordon-Quayle K, Gorini G, Grab J, Gracie L, Greenland M, Greenwood N, Greffrath J, Groenewald MM, Grossi L, Gupta G, Hackett M, Hallis B, Hamaluba M, Hamilton E, Hamlyn J, Hammersley D, Hanrath AT, Hanumunthadu B, Harris SA, Harris C, Harris T, Harrison TD, Harrison D, Hart TC, Hartnell B, Hassan S, Haughney J, Hawkins S, Hay J, Head I, Henry J, Hermosin Herrera M, Hettle DB, Hill J, Hodges G, Horne E, Hou MM, Houlihan C, Howe E, Howell N, Humphreys J, Humphries HE, Hurley K, Huson C, Hyder-Wright A, Hyams C, Ikram S, Ishwarbhai A, Ivan M, Iveson P, Iyer V, Jackson F, De Jager J, Jaumdally S, Jeffers H, Jesudason N, Jones B, Jones K, Jones E, Jones C, Jorge MR, Jose A, Joshi A, Júnior EAMS, Kadziola J, Kailath R, Kana F, Karampatsas K, Kasanyinga M, Keen J, Kelly EJ, Kelly DM, Kelly D, Kelly S, Kerr D, Kfouri R de Á, Khan L, Khozoee B, Kidd S, Killen A, Kinch J, Kinch P, King LDW, King TB, Kingham L, Klenerman P, Knapper F, Knight JC, Knott D, Koleva S, Lang M, Lang G, Larkworthy CW, Larwood JPJ, Law R, Lazarus EM, Leach A, Lees EA, Lemm N-M, Lessa A, Leung S, Li Y, Lias AM, Liatsikos K, Linder A, Lipworth S, Liu S, Liu X, Lloyd A, Lloyd S, Loew L, Lopez Ramon R, Lora L, Lowthorpe V, Luz K, MacDonald JC, MacGregor G, Madhavan M, Mainwaring DO, Makambwa E, Makinson R, Malahleha M, Malamatsho R, Mallett G, Mansatta K, Maoko T, Mapetla K, Marchevsky NG, Marinou S, Marlow E, Marques GN, Marriott P, Marshall RP, Marshall JL, Martins FJ, Masenya M, Masilela M, Masters SK, Mathew M, Matlebjane H, Matshidiso K, Mazur O, Mazzella A, McCaughan H, McEwan J, McGlashan J, McInroy L, McIntyre Z, McLenaghan D, McRobert N, McSwiggan S, Megson C, Mehdipour S, Meijs W, Mendonça RNÁ, Mentzer AJ, Mirtorabi N, Mitton C, Mnyakeni S, Moghaddas F, Molapo K, Moloi M, Moore M, Moraes-Pinto MI, Moran M, Morey E, Morgans R, Morris S, Morris S, Morris HC, Morselli F, Morshead G, Morter R, Mottal L, Moultrie A, Moya N, Mpelembue M, Msomi S, Mugodi Y, Mukhopadhyay E, Muller J, Munro A, Munro C, Murphy S, Mweu P, Myasaki CH, Naik G, Naker K, Nastouli E, Nazir A, Ndlovu B, Neffa F, Njenga C, Noal H, Noé A, Novaes G, Nugent FL, Nunes G, O’Brien K, O’Connor D, Odam M, Oelofse S, Oguti B, Olchawski V, Oldfield NJ, Oliveira MG, Oliveira C, Oosthuizen A, O’Reilly P, Osborne P, Owen DRJ, Owen L, Owens D, Owino N, Pacurar M, Paiva BVB, Palhares EMF, Palmer S, Parkinson S, Parracho Hmrt, Parsons K, Patel D, Patel B, Patel F, Patel K, Patrick-Smith M, Payne RO, Peng Y, Penn EJ, Pennington A, Peralta Alvarez MP, Perring J, Perry N, Perumal R, Petkar S, Philip T, Phillips DJ, Phillips J, Phohu MK, Pickup L, Pieterse S, Piper J, Pipini D, Plank M, Du Plessis J, Pollard S, Pooley J, Pooran A, Poulton I, Powers C, Presa FB, Price DA, Price V, Primeira M, Proud PC, Provstgaard-Morys S, Pueschel S, Pulido D, Quaid S, Rabara R, Radford A, Radia K, Rajapaska D, Rajeswaran T, Ramos ASF, Ramos Lopez F, Rampling T, Rand J, Ratcliffe H, Rawlinson T, Rea D, Rees B, Reiné J, Resuello-Dauti M, Reyes Pabon E, Ribiero CM, Ricamara M, Richter A, Ritchie N, Ritchie AJ, Robbins AJ, Roberts H, Robinson RE, Robinson H, Rocchetti TT, Rocha BP, Roche S, Rollier C, Rose L, Ross Russell AL, Rossouw L, Royal S, Rudiansyah I, Ruiz S, Saich S, Sala C, Sale J, Salman AM, Salvador N, Salvador S, Sampaio M, Samson AD, Sanchez-Gonzalez A, Sanders H, Sanders K, Santos E, Santos Guerra Mfs, Satti I, Saunders JE, Saunders C, Sayed A, Schim van der Loeff I, Schmid AB, Schofield E, Screaton G, Seddiqi S, Segireddy RR, Senger R, Serrano S, Shah R, Shaik I, Sharpe HE, Sharrocks K, Shaw R, Shea A, Shepherd A, Shepherd JG, Shiham F, Sidhom E, Silk SE, da Silva Moraes AC, Silva-Junior G, Silva-Reyes L, Silveira AD, Silveira MBV, Sinha J, Skelly DT, Smith DC, Smith N, Smith HE, Smith DJ, Smith CC, Soares A, Soares T, Solórzano C, Sorio GL, Sorley K, Sosa-Rodriguez T, Souza CMCDL, Souza BSDF, Souza AR, Spencer AJ, Spina F, Spoors L, Stafford L, Stamford I, Starinskij I, Stein R, Steven J, Stockdale L, Stockwell LV, Strickland LH, Stuart AC, Sturdy A, Sutton N, Szigeti A, Tahiri-Alaoui A, Tanner R, Taoushanis C, Tarr AW, Taylor K, Taylor U, Taylor IJ, Taylor J, te Water Naude R, Themistocleous Y, Themistocleous A, Thomas M, Thomas K, Thomas TM, Thombrayil A, Thompson F, Thompson A, Thompson K, Thompson A, Thomson J, Thornton-Jones V, Tighe PJ, Tinoco LA, Tiongson G, Tladinyane B, Tomasicchio M, Tomic A, Tonks S, Towner J, Tran N, Tree J, Trillana G, Trinham C, Trivett R, Truby A, Tsheko BL, Turabi A, Turner R, Turner C, Ulaszewska M, Underwood BR, Varughese R, Verbart D, Verheul M, Vichos I, Vieira T, Waddington CS, Walker L, Wallis E, Wand M, Warbick D, Wardell T, Warimwe G, Warren SC, Watkins B, Watson E, Webb S, Webb-Bridges A, Webster A, Welch J, Wells J, West A, White C, White R, Williams P, Williams RL, Winslow R, Woodyer M, Worth AT, Wright D, Wroblewska M, Yao A, Zimmer R, Zizi D & Zuidewind P (2021) Safety and efficacy of the ChAdOx1 nCoV-19 vaccine (AZD1222) against SARS-CoV-2: an interim analysis of four randomised controlled trials in Brazil, South Africa, and the UK. The Lancet 397, 99–111.

19 Ella R, Vadrevu KM, Jogdand H, Prasad S, Reddy S, Sarangi V, Ganneru B, Sapkal G, Yadav P, Abraham P, Panda S, Gupta N, Reddy P, Verma S, Kumar Rai S, Singh C, Redkar SV, Gillurkar CS, Kushwaha JS, Mohapatra S, Rao V, Guleria R, Ella K & Bhargava B (2021) Safety and immunogenicity of an inactivated SARS-CoV-2 vaccine, BBV152: a double-blind, randomised, phase 1 trial. The Lancet Infectious Diseases 21, 637–646.

20 Elbe S & Buckland-Merrett G (2017) Data, disease and diplomacy: GISAID’s innovative contribution to global health: Data, Disease and Diplomacy. Global Challenges 1, 33–46.

21 Bolger AM, Lohse M & Usadel B (2014) Trimmomatic: a flexible trimmer for Illumina sequence data. Bioinformatics 30, 2114–2120.

22 Li H & Durbin R (2009) Fast and accurate short read alignment with Burrows-Wheeler transform. Bioinformatics 25, 1754–1760.

23 Li H, Handsaker B, Wysoker A, Fennell T, Ruan J, Homer N, Marth G, Abecasis G, Durbin R, & 1000 Genome Project Data Processing Subgroup (2009) The Sequence Alignment/Map format and SAMtools. Bioinformatics 25, 2078–2079.

24 Grubaugh ND, Gangavarapu K, Quick J, Matteson NL, De Jesus JG, Main BJ, Tan AL, Paul LM, Brackney DE, Grewal S, Gurfield N, Van Rompay KKA, Isern S, Michael SF, Coffey LL, Loman NJ & Andersen KG (2019) An amplicon-based sequencing framework for accurately measuring intrahost virus diversity using PrimalSeq and iVar. Genome Biol 20, 8.

25 Cingolani P, Platts A, Wang LL, Coon M, Nguyen T, Wang L, Land SJ, L. X & Ruden DM (2012) A program for annotating and predicting the effects of single nucleotide polymorphisms, SnpEff: SNPs in the genome of Drosophila melanogaster strain w1118; iso-2; iso-3. Fly (Austin) 6, 80–92.

26 Rambaut A, Holmes EC, O’Toole Á, Hill V, McCrone JT, Ruis C, du Plessis L & Pybus OG (2020) A dynamic nomenclature proposal for SARS-CoV-2 lineages to assist genomic epidemiology. Nat Microbiol 5, 1403–1407.

27 Friendly M (2002) Corrgrams: Exploratory Displays for Correlation Matrices. The American Statistician 56, 316–324.

28 Wilm A, Aw PPK, Bertrand D, Yeo GHT, Ong SH, Wong CH, Khor CC, Petric R, Hibberd ML & Nagarajan N (2012) LoFreq: a sequence-quality aware, ultra-sensitive variant caller for uncovering cell-population heterogeneity from high-throughput sequencing datasets. Nucleic Acids Res 40, 11189–11201.

29 O’Toole Á, Scher E, Underwood A, Jackson B, Hill V, McCrone JT, Colquhoun R, Ruis C, Abu-Dahab K, Taylor B, Yeats C, Du Plessis L, Maloney D, Medd N, Attwood SW, Aanensen DM, Holmes EC, Pybus OG & Rambaut A (2021) Assignment of Epidemiological Lineages in an Emerging Pandemic Using the Pangolin Tool. Virus Evolution, veab064.

30 Singh J, Rahman SA, Ehtesham NZ, Hira S & Hasnain SE (2021) SARS-CoV-2 variants of concern are emerging in India. Nat Med 27, 1131–1133.

31 Tonkin-Hill G, Martincorena I, Amato R, Lawson AR, Gerstung M, Johnston I, Jackson DK, Park N, Lensing SV, Quail MA, Gonçalves S, Ariani C, Spencer Chapman M, Hamilton WL, Meredith LW, Hall G, Jahun AS, Chaudhry Y, Hosmillo M, Pinckert ML, Georgana I, Yakovleva A, Caller LG, Caddy SL, Feltwell T, Khokhar FA, Houldcroft CJ, Curran MD, Parmar S, COVID-19 Genomics UK (COG-UK) Consortium, Alderton A, Nelson R, Harrison EM, Sillitoe J, Bentley SD, Barrett JC, Torok ME, Goodfellow IG, Langford C, Kwiatkowski D, & Wellcome Sanger Institute COVID-19 Surveillance Team (2021) Patterns of within-host genetic diversity in SARS-CoV-2. Elife 10, e66857.

32 Jackson B, Boni MF, Bull MJ, Colleran A, Colquhoun RM, Darby AC, Haldenby S, Hill V, Lucaci A, McCrone JT, Nicholls SM, O’Toole Á, Pacchiarini N, Poplawski R, Scher E, Todd F, Webster HJ, Whitehead M, Wierzbicki C, Loman NJ, Connor TR, Robertson DL, Pybus OG & Rambaut A (2021) Generation and transmission of interlineage recombinants in the SARS-CoV-2 pandemic. Cell 184, 5179–5188.e8.

33 Valesano AL, Rumfelt KE, Dimcheff DE, Blair CN, Fitzsimmons WJ, Petrie JG, Martin ET & Lauring AS (2021) Temporal dynamics of SARS-CoV-2 mutation accumulation within and across infected hosts. PLoS Pathog 17, e1009499.

34 Lythgoe KA, Hall M, Ferretti L, de Cesare M, MacIntyre-Cockett G, Trebes A, Andersson M, Otecko N, Wise EL, Moore N, Lynch J, Kidd S, Cortes N, Mori M, Williams R, Vernet G, Justice A, Green A, Nicholls SM, Ansari MA, Abeler-Dörner L, Moore CE, Peto TEA, Eyre DW, Shaw R, Simmonds P, Buck D, Todd JA, Connor TR, da Silva Filipe A, Shepherd J, Thomson EC, The COVID-19 Genomics UK (COG-UK) consortium, Bonsall D, Fraser C & Golubchik T (2020) Within-host genomics of SARS-CoV-2 Genomics.

35 Martin MA, VanInsberghe D & Koelle K (2021) Insights from SARS-CoV-2 sequences. Science 371, 466–467.

36 Ovsyannikova IG, Haralambieva IH, Crooke SN, Poland GA & Kennedy RB (2020) The role of host genetics in the immune response to SARS-CoV-2 and COVID-19 susceptibility and severity. Immunol Rev 296, 205–219.

37 Di Maria E, Latini A, Borgiani P & Novelli G (2020) Genetic variants of the human host influencing the coronavirus-associated phenotypes (SARS, MERS and COVID-19): rapid systematic review and field synopsis. Hum Genomics 14, 30.

38 Andreano E, Piccini G, Licastro D, Casalino L, Johnson NV, Paciello I, Dal Monego S, Pantano E, Manganaro N, Manenti A, Manna R, Casa E, Hyseni I, Benincasa L, Montomoli E, Amaro RE, McLellan JS & Rappuoli R (2021) SARS-CoV-2 escape from a highly neutralizing COVID-19 convalescent plasma. Proc Natl Acad Sci USA 118, e2103154118.

39 Dhar MS, Marwal R, Vs R, Ponnusamy K, Jolly B, Bhoyar RC, Sardana V, Naushin S, Rophina M, Mellan TA, Mishra S, Whittaker C, Fatihi S, Datta M, Singh P, Sharma U, Ujjainiya R, Bhatheja N, Divakar MK, Singh MK, Imran M, Senthivel V, Maurya R, Jha N, Mehta P A V, Sharma P, Vr A, Chaudhary U, Soni N, Thukral L, Flaxman S, Bhatt S, Pandey R, Dash D, Faruq M, Lall H, Gogia H, Madan P, Kulkarni S, Chauhan H, Sengupta S, Kabra S, The Indian SARS-CoV-2 Genomics Consortium (INSACOG)‡, Gupta RK, Singh SK, Agrawal A, Rakshit P, Nandicoori V, Tallapaka KB, Sowpati DT, Thangaraj K, Bashyam MD, Dalal A, Sivasubbu S, Scaria V, Parida A, Raghav SK, Prasad P, Sarin A, Mayor S, Ramakrishnan U, Palakodeti D, Seshasayee ASN, Bhat M, Shouche Y, Pillai A, Dikid T, Das S, Maitra A, Chinnaswamy S, Biswas NK, Desai AS, Pattabiraman C, Manjunatha MV, Mani RS, Arunachal Udupi G, Abraham P, Atul PV & Cherian SS (2021) Genomic characterization and epidemiology of an emerging SARS-CoV-2 variant in Delhi, India. Science, eabj9932.

40 Singanayagam A, Hakki S, Dunning J, Madon KJ, Crone MA, Koycheva A, Derqui-Fernandez N, Barnett JL, Whitfield MG, Varro R, Charlett A, Kundu R, Fenn J, Cutajar J, Quinn V, Conibear E, Barclay W, Freemont PS, Taylor GP, Ahmad S, Zambon M, Ferguson NM, Lalvani A, Badhan A, Dustan S, Tejpal C, Ketkar AV, Narean JS, Hammett S, McDermott E, Pillay T, Houston H, Luca C, Samuel J, Bremang S, Evetts S, Poh J, Anderson C, Jackson D, Miah S, Ellis J & Lackenby A (2021) Community transmission and viral load kinetics of the SARS-CoV-2 delta (B.1.617.2) variant in vaccinated and unvaccinated individuals in the UK: a prospective, longitudinal, cohort study. The Lancet Infectious Diseases, S1473309921006484.

41 Li B, Deng A, Li K, Hu Y, Li Z, Xiong Q, Liu Z, Guo Q, Zou L, Zhang H, Zhang M, Ouyang F, Su J, Su W, Xu J, Lin H, Sun J, Peng J, Jiang H, Zhou P, Hu T, Luo M, Zhang Y, Zheng H, Xiao J, Liu T, Che R, Zeng H, Zheng Z, Huang Y, Yu J, Yi L, Wu J, Chen J, Zhong H, Deng X, Kang M, Pybus OG, Hall M, Lythgoe KA, Li Y, Yuan J, He J & Lu J (2021) Viral infection and transmission in a large, well-traced outbreak caused by the SARS-CoV-2 Delta variant Epidemiology.

42 Grenfell BT, Pybus OG, Gog JR, Wood JLN, Daly JM, Mumford JA & Holmes EC (2004) Unifying the Epidemiological and Evolutionary Dynamics of Pathogens. Science 303, 327–332.

43 McCallum M, Walls AC, Sprouse KR, Bowen JE, Rosen LE, Dang HV, De Marco A, Franko N, Tilles SW, Logue J, Miranda MC, Ahlrichs M, Carter L, Snell G, Pizzuto MS, Chu HY, Van Voorhis WC, Corti D & Veesler D (2021) Molecular basis of immune evasion by the Delta and Kappa SARS-CoV-2 variants. Science 374, 1621–1626.

44 Karamitros T, Papadopoulou G, Bousali M, Mexias A, Tsiodras S & Mentis A (2020) SARS-CoV-2 exhibits intra-host genomic plasticity and low-frequency polymorphic quasispecies. Journal of Clinical Virology 131, 104585.

45 Wang Y, Wang D, Zhang L, Sun W, Zhang Z, Chen W, Zhu A, Huang Y, Xiao F, Yao J, Gan M, Li F, Luo L, Huang X, Zhang Y, Wong S, Cheng X, Ji J, Ou Z, Xiao M, Li M, Li J, Ren P, Deng Z, Zhong H, Xu X, Song T, Mok CKP, Peiris M, Zhong N, Zhao J, Li Y, Li J & Zhao J (2021) Intra-host variation and evolutionary dynamics of SARS-CoV-2 populations in COVID-19 patients. Genome Med 13, 30.

46 Lythgoe KA, Hall M, Ferretti L, de Cesare M, MacIntyre-Cockett G, Trebes A, Andersson M, Otecko N, Wise EL, Moore N, Lynch J, Kidd S, Cortes N, Mori M, Williams R, Vernet G, Justice A, Green A, Nicholls SM, Ansari MA, Abeler-Dörner L, Moore CE, Peto TEA, Eyre DW, Shaw R, Simmonds P, Buck D, Todd JA, on behalf of the Oxford Virus Sequencing Analysis Group (OVSG)‡, Connor TR, Ashraf S, da Silva Filipe A, Shepherd J, Thomson EC, The COVID-19 Genomics UK (COG-UK) Consortium§, Bonsall D, Fraser C & Golubchik T (2021) SARS-CoV-2 within-host diversity and transmission. Science 372, eabg0821.

47 Bianchi M, Borsetti A, Ciccozzi M & Pascarella S (2021) SARS-Cov-2 ORF3a: Mutability and function. Int J Biol Macromol 170, 820–826.

48 Issa E, Merhi G, Panossian B, Salloum T & Tokajian S (2020) SARS-CoV-2 and ORF3a: Nonsynonymous Mutations, Functional Domains, and Viral Pathogenesis. mSystems 5, e00266–20, /msystems/5/3/msys.00266-20.atom.

49 Ren Y, Shu T, Wu D, Mu J, Wang C, Huang M, Han Y, Zhang X-Y, Zhou W, Qiu Y & Zhou X (2020) The ORF3a protein of SARS-CoV-2 induces apoptosis in cells. Cell Mol Immunol 17, 881–883.

50 Bascos NAD, Mirano-Bascos D & Saloma CP (2021) Structural Analysis of Spike Protein Mutations in the SARS-CoV-2 P.3 Variant Biophysics.

51 Chand GB, Banerjee A & Azad GK (2020) Identification of twenty-five mutations in surface glycoprotein (Spike) of SARS-CoV-2 among Indian isolates and their impact on protein dynamics. Gene Reports 21, 100891.

